# Hindsight is 2020 vision: a characterisation of the global response to the COVID-19 pandemic

**DOI:** 10.1101/2020.04.30.20085662

**Authors:** David J. Warne, Anthony Ebert, Christopher Drovandi, Wenbiao Hu, Antonietta Mira, Kerrie Mengersen

## Abstract

**Background:** The global impact of COVID-19 and the country-specific responses to the pandemic provide an unparalleled opportunity to learn about different patterns of the outbreak and interventions. We model the global pattern of trajectories of reported COVID-19 cases during the primary response period, with the aim of learning from the past to prepare for the future.

**Methods:** Using Bayesian methods, we analyse the response to the COVID-19 outbreak for 158 countries for the period 22 January to 9 June 2020. This encompasses the period in which many countries imposed a variety of response measures and initial relaxation strategies. Instead of modelling specific intervention types and timings for each country explicitly, we adopt a stochastic epidemiological model including a feedback mechanism on virus transmission to capture complex nonlinear dynamics arising from continuous changes in community behaviour in response to rising case numbers. We analyse the overall effect of interventions and community responses across diverse regions. This approach mitigates explicit consideration of issues such as period of infectivity and public adherence to government restrictions.

**Results:** Countries with the largest cumulative case tallies are characterised by a delayed response, whereas countries that avoid substantial community transmission during the period of study responded quickly. Countries that recovered rapidly also have a higher case identification rate and small numbers of undocumented community transmission at the early stages of the outbreak. We also demonstrate that uncertainty in numbers of undocumented infections dramatically impacts the risk of second waves. Our approach is also effective at pre-empting potential second waves and flare-ups.

**Conclusions:** We demonstrate the utility of modelling to interpret community behaviour in the early epidemic stages. Two lessons learnt that are important for the future are: i) countries that imposed strict containment measures early in the epidemic fared better with respect to numbers of reported cases; and ii) broader testing is required early in the epidemic to understand the magnitude of undocumented infections and recover rapidly. We conclude that clear patterns of containment are essential prior to relaxation of restrictions and show that modelling can provide insights to this end.

## Background

The world is at war with the coronavirus disease 2019 (COVID-19) [1], which was officially declared a pandemic by the World Health Organization (WHO) on 11 March 2020 [2]. From early cases reported in December 2019 (Hubei, China), severe acute respiratory syndrome coronavirus 2 (SARS-CoV-2) spread rapidly throughout the world [3,4]. The challenge faced by healthcare systems worldwide cannot be understated [5-8]. Within four months of the first reported cases, more than two and a half million cases were confirmed with over two hundred thousand deaths globally [9,10], and most countries had taken a range of extreme measures to stop the spread. More than six months on, the pandemic is far from over but many countries have cautiously started to relax restrictions. A unique feature of the COVID-19 pandemic has been the rapid and widespread availability of data though online platforms [11-13]. These data enable the analysis of various patterns of outbreak containment over this period, and provide an unparalleled opportunity to learn how we might respond to a potential second surge of COVID-19 or future pandemics. To this end, it is important to learn from the past in order to prepare for the future.

Travel restrictions, increased hygiene education, social distancing, school and business closures, and complete lockdowns [1,8,14] are examples of non-pharmaceutical intervention (NPI) strategies that many countries have introduced to slow transmission rates and relieve pressure on healthcare systems in the absence of a vaccine or treatment for COVID-19 [15]. Modelling and simulation are at the forefront of determining the efficacy of these measures in reducing SARS-CoV-2 transmission and quantifying risk of future outbreaks along with their potential severity [16-19]. Understanding of these effects is crucial given the potential deleterious sociological and economic impacts of many NPIs [1,20-22].

The global modelling community has responded rapidly to the COVID-19 situation and has provided insight into the transmissibility of SARS-CoV-2 [23-25], global risk of spread through transport networks [10,26], forecasting and prediction [27-29], and evaluation of interventions [18,30]. A variety of modelling strategies have been applied. Techniques include: empirical approaches such as phenomenological growth curves [29]; data-driven, statistical approaches using non-linear autoregressive models [31]; and mechanistic models based on epidemiological theory [32] with various extensions [33,34].

Our aim is to learn about the global pattern of behaviour among countries based on the trajectories of reported cases, recoveries and fatalities as provided by Johns Hopkins University [9,10]. Although there are acknowledged drawbacks in relying on reported cases, we argue that such data will be the main source of information for government and health managers in future scenarios. In light of our aim, we also avoid imposing the specific complex history of intervention measures for each country, but instead include a novel regulatory mechanism that captures the changes in community behaviour in response to rising confirmed cases. This is achieved by including a feedback loop in the transmission process which enables complex nonlinear dynamics arising form continuous changes in community responses over time. Through model calibration, we are able to infer the country specific response timing and strength. As a result, we are able to analyse the overall effect of interventions and community responses across diverse regions. This approach also mitigates explicit consideration of issues such as period of infectivity and public adherence to government restrictions.

Within a Bayesian framework, we characterise the response of 158 countries to the COVID-19 outbreak for the period 22 January to 9 June 2020. We focus on this timeframe since it encompasses the period during which initial measures were imposed by these countries and excludes the period in which countries started to relax restrictions. Our characterisation is a broad assessment of the global response to the COVID-19 pandemic, and reflects how countries in early phases of outbreak may have adjusted their strategies to reduce the time to recovery. In particular, we find that very large outbreaks are characterised by a delayed response to rising confirmed case numbers before significant reductions in transmission rates occur. Countries that observed a decrease in active cases during the early period of study (e.g., China and South Korea) are characterised by high case identification rates; the result of highly effective identification and quarantine programs. We also find that, for many countries, the transmission rates in the later period of the study are declining. However, large unobserved infected population counts are also estimated. Our analysis confirms that a multifaceted approach that includes NPIs, increased testing, contact tracing, isolation and quarantine measures are effective in reducing the severity of COVID-19 outbreaks world-wide. We also demonstrate, using data up to 9 June 2020, that the unknown magnitude of undocumented cases substantively impacts uncertainty in risks of subsequent increases after recovery. We conclude that wider testing is also essential to reduce this uncertainty in the asymptomatic infected populations to reliably evaluate risk of second waves of COVID-19. This is essential as restrictions are eased around the world.

## Methods

### Data summary

Daily counts of reported confirmed COVID-19 cases, recoveries and deaths for each country are obtained from the Johns Hopkins University coronavirus resource center [11]. We refer the reader to the Discussion section for comments on this data source. Population data for each country for 2020 were obtained from United Nations Population Division estimates [35].

Our analysis is performed over three different time periods: i) 22 January-30 March; and ii) 22 January-13 April; and iii) 22 January-9 June. These periods are selected as they broadly represent the time period of the initial outbreak of COVID-19; covering the initial exponential growth period, the epidemic peak, and subsequent recovery period of many countries worldwide. In particular, we use these time points to look at the changes in key model parameters relating to a countries responses over time.

Countries are included in the analysis for a give time period provided the cumulative number of confirmed COVID-19 cases exceeded 100 at least one day prior to the end of the particular analysis period. This condition is enforced to ensure sufficient mixing and that enough days of daily counts are observed for sensible model fitting and parameter inferences. Using these inclusion criteria, we obtain *N* = 98 countries for the period of 22 January-30 March, *N* = 121 countries for period of 22 January to 13 April, and *N* = 158 countries for the period 22 January-9 June.

### Analysis summary

For each country, *i* = 1, 2,…, *N*, the Johns Hopkins University maintains a time-series, 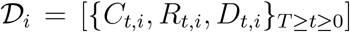, where *C_t_,_i_, R_t,i_*, and *D_t_,_i_* are, respectively, the cumulative confirmed cases, case recoveries and case deaths on day t for country *i, t* = 0 is the first day such that *C_t,i_* ≥ 100 and *t* = *T* is the end of the study period. Since it is known that there are variations in reporting protocols across countries and time as well as data curation challenges [12], caution is necessary in the interpretation of our analysis across all countries over time.

Bayesian parameter inference is applied over three time periods. The first period, 22 January to 30 March, is used to assess the community response to the initial outbreak of COVID-19. This time period is chosen as it reflects the period when many countries had not yet introduced any interventions, and those that did introduce such measures had not yet seen the effect of them. Our analysis for this period represents a reflection on how countries initially responded to the pandemic.

Our second analysis period, including data up to 13 April, encompasses the time period in which the efficacy of the community response starts to become evident. For several countries, a decline in daily cases is observed. Therefore, this period allows us to investigate how different strategies impacted the overall cumulative case numbers and the time to recovery.

Our third analysis considers the period up to the 9 June, in which many countries had started to relax restrictions. Furthermore, the epicentre of the pandemic had shifted from Europe to the Americas. We note the shift in parameter estimates for recovering European countries and those countries of South America that were observing exponential growth in daily cases. We also consider in this analysis the prevention of second waves, and highlight the sensitivity of the system dynamics to the uncertainty in unobserved infectious individuals.

### Mathematical model

Our model is a stochastic epidemiological compartmental model that describes the spread of COVID-19 within a single country over the time period *t* ∊ (0, *T*]. The assumed well-mixed population of size *P* is comprised of six compartments: susceptible, *S_t_;* infectious, *I_t_*; confirmed active cases, *A_t_;* case recoveries, *R_t_*; case fatalities, *D_t_*; and unconfirmed recoveries, *R_t_*; Here, the population that is susceptible to the SARS-CoV-2 infection, *S_t_*, can be infected by infectious individuals, *I_t_*. Importantly, *I_t_* represents the unobserved infectious population, including both symptomatic and asymptomatic infections. The active confirmed cases, *A_t_*, are those who have tested positive for COVID-19 but have not yet recovered or died. It is assumed that individuals in *A_t_* are isolated from the susceptible community (e.g., self-isolated, quarantined, or hospitalised) and no longer contribute to new infections; importantly, *A_t_* need not be symptomatic, but may have been identified as a result of contact tracing protocols or community wide testing. *R_t_* and *D_t_* are, respectively, the population of confirmed cases that recover or die; these sub-populations correspond to the recoveries and deaths reported in the Johns Hopkins University data. Lastly, RU is the population of infected individuals that recover or die without being tested for COVID-19; these individuals no longer spread the infection but do not contribute to the reported recovery and fatality counts. The cumulative confirmed cases, as reported by the Johns Hopkins University, can be obtained by *C_t_* = *A_t_ + R_t_ + D_t_*.

The true values of *S_t_, I_t_*, and 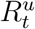 are not observable in reality and are latent variables in our model. As a result, strategies for managing the spread of the virus, such as NPIs, are informed by the observed populations *A_t_, R_t_*, and *D_t_*. Furthermore, media coverage, official government information, and health authority reports based on these observed quantities may also affect the behaviour of individuals. For example, frequent reports on growing case numbers may increase voluntary self-isolation; conversely, media coverage that downplays the risk of infection may lead to widespread non-compliance with health advice or government regulations. We model this dynamic by treating the transmission rate as a non-linear function of the observable populations, *g*(*A_t_, R_t_, D_t_*), thus introducing a feedback loop.

A schematic of this system that highlights the state transitions and the feedback loop is given in Fig. 1. The resulting dynamics can be described by the differential equations,

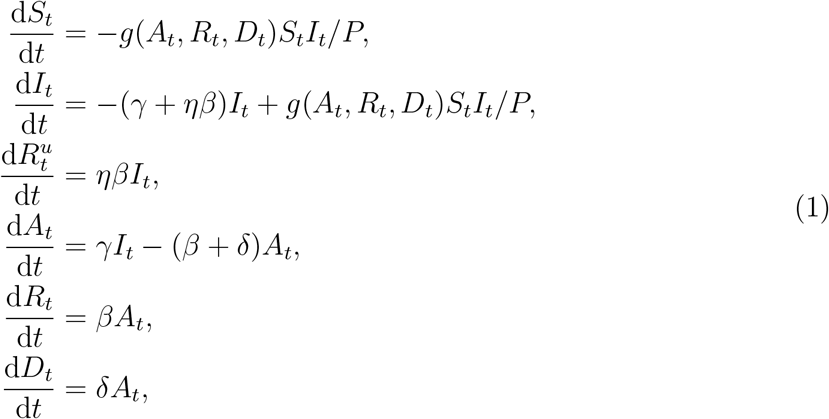

d*t* where *g*(·) *>* 0 is the non-linear transmission rate function, *γ>* 0 is the identification rate, *β>* 0 is the case recovery rate, *δ>* 0 is the case fatality rate, and *η>* 0 is the latent removal rate relative to the case recovery rate. The initial conditions for the observables, *A*_0_*,R*_0_*,D*_0_, are obtained from the Johns Hopkins University data. To capture uncertainty in community spread at early time we set the initial infectious population to *I*_0_ = *κA*_0_, where *κ>* 0 is the relative number of unobserved cases. Finally, we assume initially 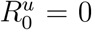, hence *S*_0_ = *P* − *C*_0_ − *I*_0_. Although we present Equation (1) as a deterministic system for ease of interpretation, we apply a stochastic equivalent that is a discrete-state continuous-time Markov process. The tau-leaping stochastic simulation scheme [36] is applied to generate approximate sample-paths of this model. See the Supplementary Material for the full stochastic formulation. The novel component of our approach is the feedback mechanism that provides a general framework to describe how communities change their behaviour as case numbers rise. This is similar to the influence of media reports that have been the subject of study for other infections diseases, such as influenza and HIV [37, 38]. However, our approach includes a parameter governing the strength of the response.

**Figure 1:**
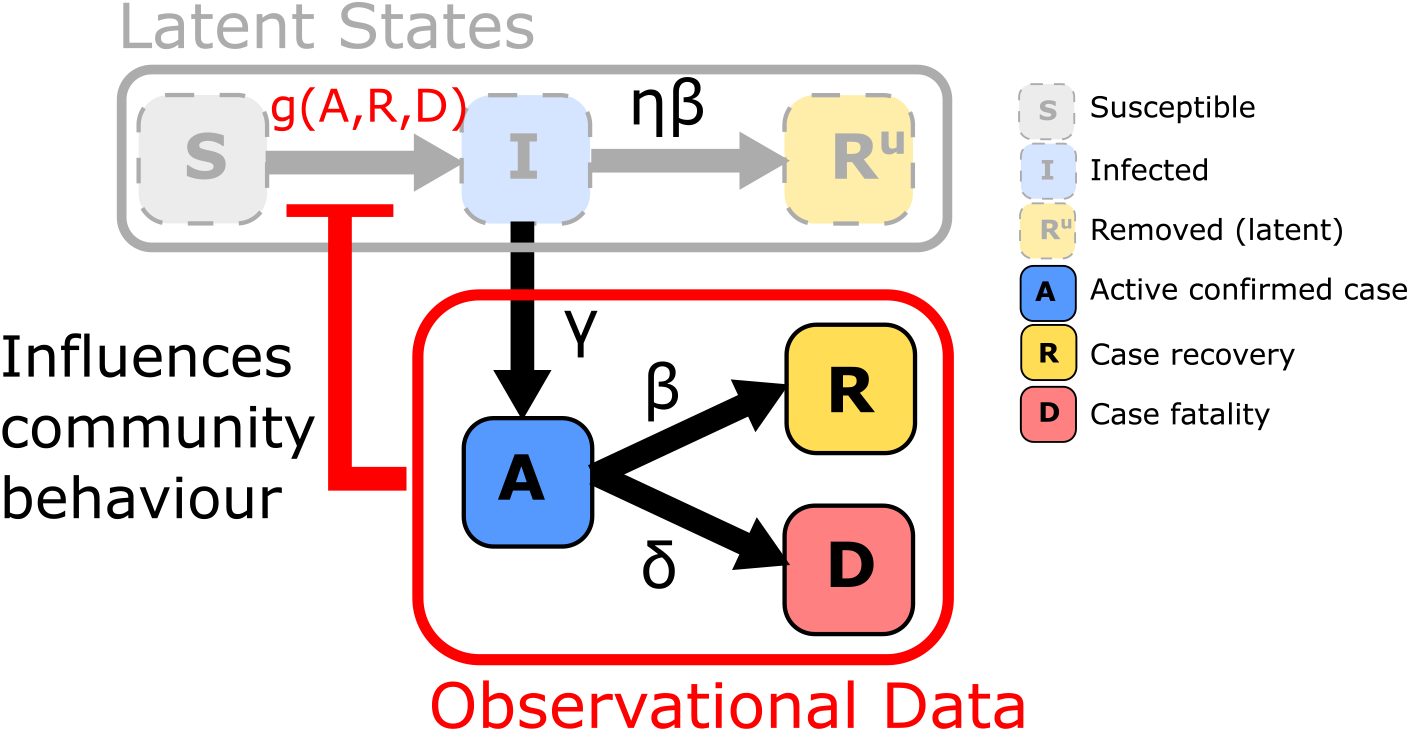
Schematic of epidemic model including a regulatory mechanism inducing a feedback loop. State transitions are marked by arrows with superscripts indicating respective rate parameters. Here, observable quantities can inform individual behaviour to inhibit transmission in the latent SIR model.

We define a reporting function,

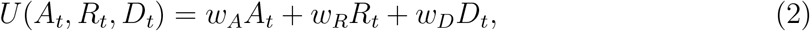

where the weights *w_A_,w_R_,w_D_* ≥ 0, represent the relative weighting of the COVID-19 data in contributing to information that influences individual behaviour, introduction of NPIs, and subsequent compliance with government regulation or health advice. In the context of this work, the weights *w_A_,w_R_*, and *w_D_* have a very important interpretation, but we first need to present more details of the feedback mechanism.

We consider a nonlinear transmission rate of the form,

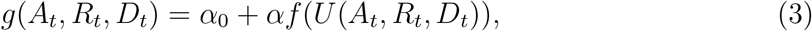

where the response function, *f*(·) ∈ [0, 1], is a decreasing function with respect to *U*(·), *α* is the controllable transmission rate such that *αf*(·) is a transmission rate that decreases as the reporting function increases and *α*_0_ is the residual transmission rate as *f*(·) → 0. Note that *α*_0_ *<g*(·) ≤ *α*_0_ + *α*. The strength of the response *s* = (1 − *f*(·)) × 100% is the percentage reduction in community transmission, excluding residual transmission *α*_0_.

For the response function we assume the form

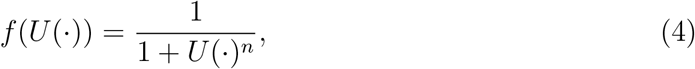

where the parameter *n* ≥ 0 controls the rate of decrease with respect to the reporting function. This form is selected for two reasons. Firstly, it is a generalisation of techniques that capture the influence of media reports during epidemics [37]. Secondly, the weights from Equation (2) have an important interpretation. This can been seen by noting that values for *A_t_,R_t_* and *D_t_* that satisfy the condition *U*(*A_t_,R_t_,D_t_*) = 1, indicate the threshold case numbers that leads to a response strength of 50%, that is, *f*(*U*(·)) = 1*/*2 leading to *g*(·)= *α*_0_ + *α/*2. The effect of varying slope parameter, *n*, and the weights are shown in Fig. 2.

**Figure 2:**
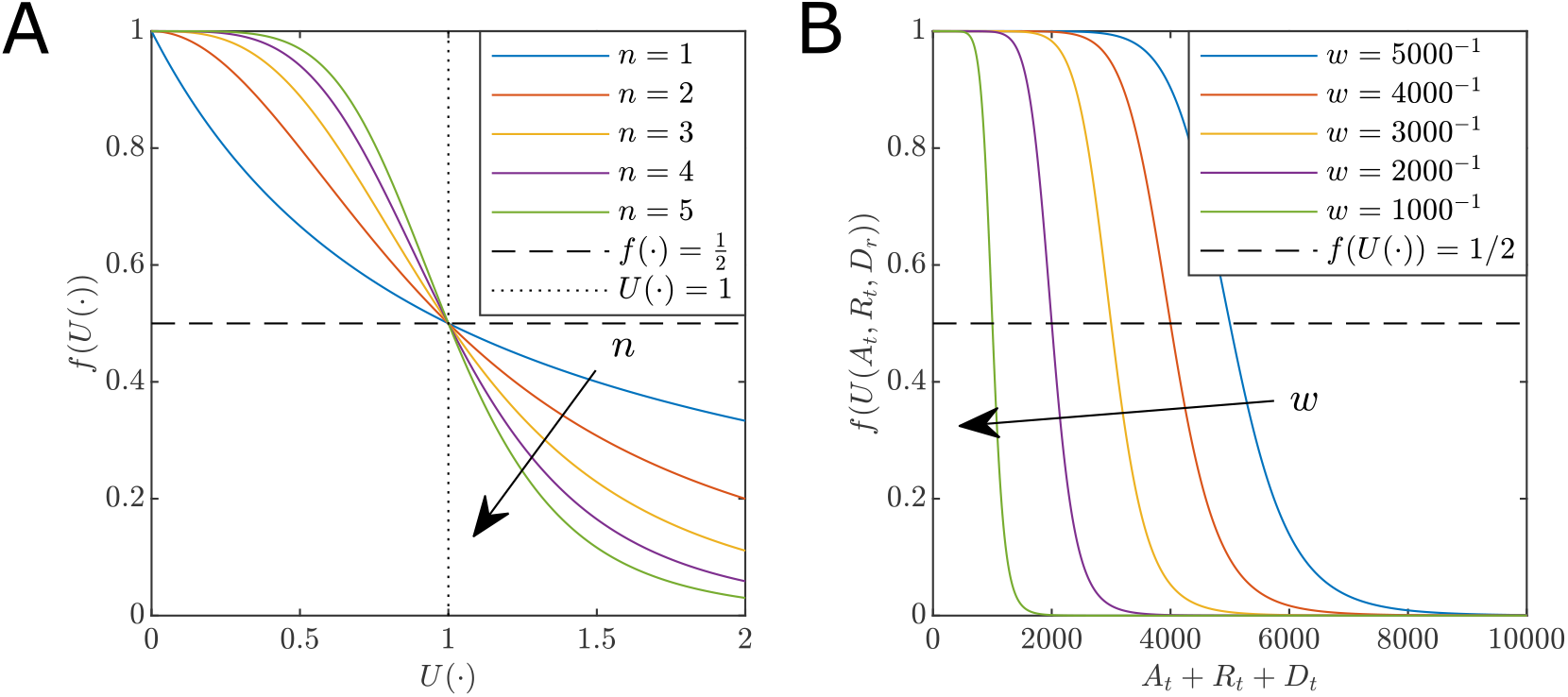
Effect of parameters on the response function. (A) The effect of the slope parameter *n*. Note, as *n* increases, the faster *f*(·) → 0. For any *n* we have *f*(·)=1*/*2 (dashed black), at the point *U*(·) = 1 (dotted black). (B) The effect of weights on the response function for the special case *w_A_* = *w_R_* = *w_D_* = *w>* 0 for constant *n* = 5. Note the point at which *f*(·)=1*/*2 corresponds to *A_t_* + *R_t_* + *D_t_* =1*/w*. That is, as *w* increases the lower the number of cases are required to influence the community to reduce the spread.

Furthermore, if *U*(·) = 0, that is no cases are reported (or the weights *w_A_* = *w_R_* = *w_D_* = 0, indicating the community does not perceive any risk), then the model reduces to a standard SIR model in the unobserved population with transmission rate parameter *α*_0_ + *α*.

As shown in Fig. 2(A), for lower *n* ≤ 1 the shape of *f*(*U*(·)) starts to decline rapidly leveling out. Increasing *n>* 1 results in a decreasing sigmoid curve with an inflection point at the critical values of *U*(·) = 1 in which a population response strength reaches 50%. For example, small *n* describes a population that does not significantly reduce the transmission rate until the *U*(·) is large. Conversely, larger *n* describes a population that acts decisively as a response that rapidly reduces transmission around *U*(·) = 1. Large values of weights *w_A_,w_R_,w_D_* correspond to lower acceptable thresholds of cases, including active, recovery and death counts. Lower weights lead to delayed responses. In this respect, the parameter *n* relates to the rate of intervention introduction and the weights relate to decision thresholds and subsequent compliance. Importantly, our approach does not distinguish between different NPIs and voluntary population behaviour, but rather models the net effect that reporting has on transmission rates.

When *w_A_* = *w_R_* = *w_D_* = *w>* 0, then the reporting function depends only on the cumulative incidence of COVID-19, *U*(*A_t_,R_t_,D_t_*)=(*A_t_* + *R_t_* + *D_t_*)*w* = *wC_t_*. This form of reporting function is useful for modelling the response to the initial wave of COVID-19 since it is proportional to the incidence risk for the entire population. Of course, other reporting functions could be considered. For example, since C*_t_* can only increase, the model dynamics cannot capture the effects of relaxing intervention restrictions. Therefore, for the results presented in this manuscript, we will focus on the reporting function with *w_R_* = *w_D_* = 0, that is, *U*(*A_t_,R_t_,D_t_*)= *w_A_A_t_*. Since *A_t_* will increase and decline over to course of an outbreak, the model can exhibit oscillatory behaviour that is essential for understanding the potential for second waves.

## Bayesian analysis

### Parameter inference

Depending on the choice of response function, our model has between 8 and 11 parameters to be estimated: two transmission rates, *α*_0_ and *α*, where *α* can be regulated but *α*_0_ cannot (i.e., residual unavoidable transmission); case recovery rate *β*; case identification rate *γ*; case death rate *δ*; relative latent recovery rate *η*; response slope parameter *n*; the initial infected scale factor *κ*; and the weights of the reporting function *w_A_*, *w_R_*, and *w_D_*. For this manuscript we assume w*_R_* = *w_D_* = 0 (see Supplementary Material for sensitivity analysis for the general case).

Using the COVID-19 time-series data, *D_i_*, for each country *i* ∈ [1, 2*,…,N*], we infer model parameters within a Bayesian framework. For notational convenience, we will omit the subscript *i* for the remainder of this section. We sample the joint posterior distribution for *θ* =[*α*_0_*, α, β, γ, δ, η, n, κ, w_A_*],

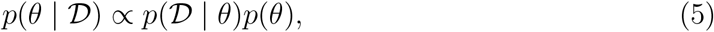

where *p*(*D*| *θ*) is the likelihood function and *p*(*θ*) is the prior distribution. To exactly evaluate the likelihood would require the solution to the Kolmogorov forward equation, which in turn requires matrix exponentiation of prohibitively large dimension. The evaluation is further complicated, as the unobserved populations *S_t_*, *I_t_*, and *R_tu_*, need to be integrated out. Therefore, the likelihood function is intractable and we rely on adaptive sequential Monte Carlo for approximate Bayesian computation (SMC-ABC) [39–42] to obtain approximate posterior samples (Supplementary Material). We use independent uniform priors, 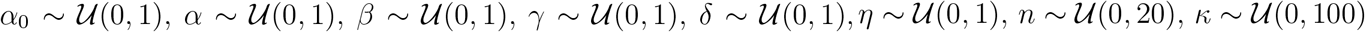, and 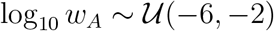.

### Assessment of model fit and prediction

The highly variable nature of the COVID-19 pandemic makes it notoriously difficult to accurately model [27, 28]. The purpose of our modelling is not to provide daily case predictions of forecasts, but rather we wish to capture the dynamic effects of changes in community behaviour during the COVID-19 outbreak. As a result, our model needs to be able to capture the overall trend in COVID-19 daily cases over time.

Model fit is assessed though sampling the posterior predictive distribution

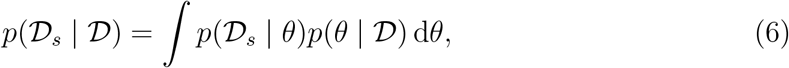

where *D_s_* is simulated data as generated by the model. We compute the 50% and 95% credible interval (CrI) of *p*(D*_s_* |D) through generating a single model simulation for each posterior sample generated by the SMC-ABC sampler and computing quantiles of the simulation state distribution at each observation time.

The posterior predictive distribution is also employed to asses the risk of second waves. After fitting the model up to 9 June, we continue to forward simulate posterior predictive samples to obtain credible intervals for oscillatory behaviour that relate to localised flare-ups and second waves.

### Parameter point estimation and uncertainty quantification

Parameter point estimates are also obtained by choosing the posterior sample that results in the lowest average discrepancy with the observed data (See Supplementary Material). Parameter uncertainty is reported using 95% CrI for the marginal approximate Bayesian posterior distributions (See Supplementary Material). We emphasise that this uncertainty quantification encompasses all plausible parameter combinations within the achieved acceptance threshold of the ABC-SMC method, rather than the average discrepancy level of the point estimate.

## Results

### Assessment of model fit

We perform a posterior predictive check to evaluate model fit (See Methods). For most countries the 95% CrI contains the daily case, recovery and death data (See Supplementary Material). Exceptions to this are usually consistent with reporting delay effects, such as weekly seasonality as evident in the daily cases for Germany (Fig. S13). While other spikes in daily numbers (e.g., Recoveries in Germany Fig. S13(H)), the model does capture overall trends well with many daily numbers remaining within the 95% credible interval. For example, the possible decline in daily cases numbers for the United States (Fig. S19(G)) is captured by the lower bound of the 95% credible interval, however, the uncertainty of this trend on 13 Apr is indicated by the increasing upper bound. In some extreme cases, such as changes in reporting methodology from China on 13 February [43](Fig. S12), subsequent consisted inaccuracies occur (Fig. S12). We discuss potential model improvements to account for this in the Discussion section. Notwithstanding this, our model appears to capture the overall trends in the trajectories to facilitate a broad comparative analysis of global responses to COVID-19 during this period.

Our approach of taking the posterior sample with the lowest expected data discrepancy is effective in this respect (See Seupplementary Material). Example model outputs using this point estimate are provided in Fig. 3 (See Supplementary Material for other examples). In particular, compare Fig. 3(A)–(B) with Fig. S19(G)–(I). Despite further increases in daily cases being highly plausible on the 13 April, we exclude these trajectories from the response analysis.

**Figure 3:**
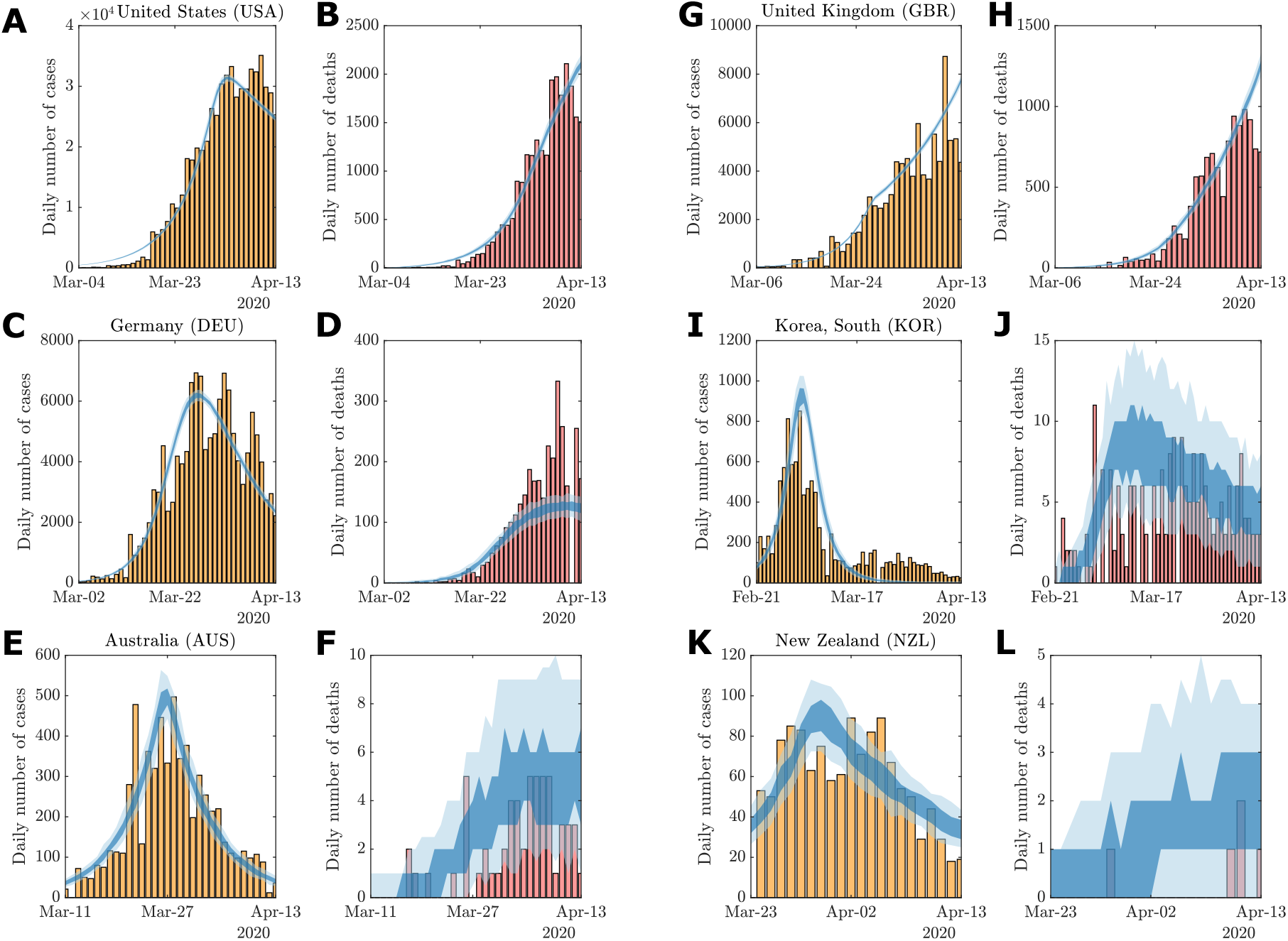
Examples of model fit using parameter point estimates: (A)–(B) United States, (C)–(D) Germany, (E)–(F) Australia, (G)–(H) United Kingdom, (I)–(J) South Korea, and (K)–(L) New Zealand. Vertical bars indicate daily reported cases (yellow) and deaths (red). The 50% (dark shaded region) and 95% credible intervals (light shaded region) of the posterior predictive distributions are plotted against the observational data. Credible intervals are computed using *n* = 100 stochastic simulations for the given point estimate. Full posterior predictive distributions are presented in the Supplementary Material.

### Characterisation of responses

We now focus on comparison of key parameters for countries of interest at each of our analysis periods. Based on our correlation analysis, these key parameters are: the case identification rate, *γ*, the relative initial undocumented infections, *κ*, and the response weight parameter *w_A_*. The fast moving evolution of epidemics and pandemics such as COVID-19 demand that model outputs and the corresponding effectiveness of mitigating measures are regularly re-assessed. Therefore, we evaluate our parameter inferences and point estimates for the three analysis periods. These results reveal that as the pandemic evolves, we can learn more about the possible response histories and latent infected populations. Figure 4 shows pairwise scatter plots for the response weight parameter *w_A_*, identification rate *γ*, and initial relative undocumented infections *κ* for each of the three analysis periods.

**Figure 4:**
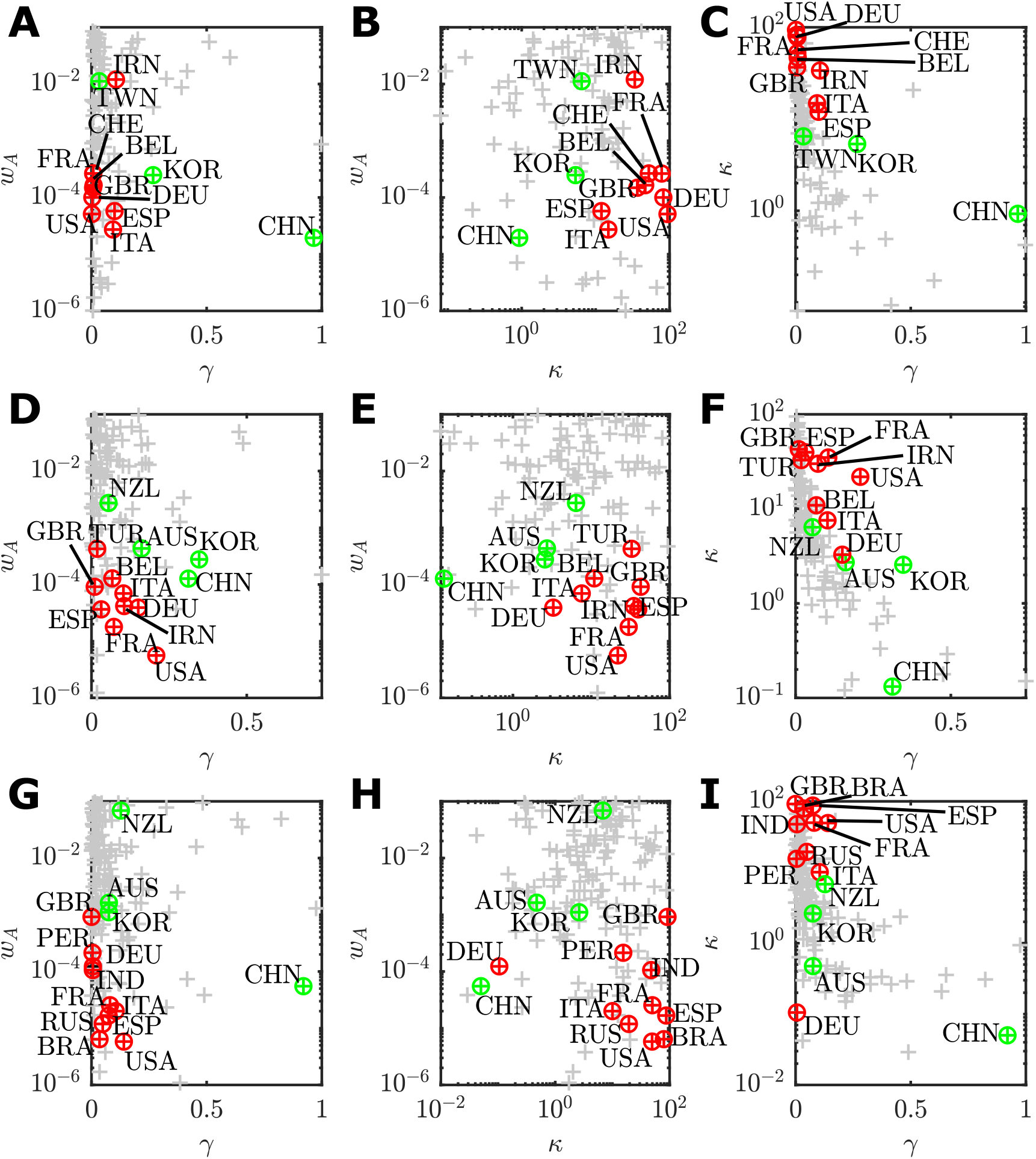
Pairwise scatter plots of point estimates of each assessed country (grey points) for the key parameters related to the management of an COVID-19 outbreak up to: (A)–(C) 30 March; (D)–(F) 13 April; and (G)–(I) 9 June. (A),(D),(G) *w_A_* versus *γ*; (B),(E),(H) *w_A_* versus *γ*; and (C),(F),(I) *κ* versus *γ*. For each time period, countries with the ten largest confirmed case counts are highlighted (red points) along with representative countries that were recovering or managed to control the outbreak (green). Labels identify the country by ISO-3166 alpha-3 code.

The interactions between parameters are complex, and the differences between time periods deserves some interpretation. However, we first highlight some overarching trends across all time periods and then discuss specific details. One clear trend is countries with that largest numbers of cumulative cases (Fig. 4, top ten countries for cumulative cases numbers indicated in red) tend to have lower response weights, typically *w_A_* ≈ 10^−4^. Some more extreme cases are as low as *w_A_* ≤ 10^−5^, for example, the United States (USA; Fig 4(D),(G)), Russia (RUS; Fig 4(G)) and Brazil (BRA; Fig 4(G)). Small *w_A_* indicates a delayed response in which the transmission rate did not decline significantly until active cases, *A_t_*, increased to larger numbers. This is consistent with reported delays in response across Europe and the United States [1]. Furthermore, low case identification rates, *γ<* 0.01, and higher relative initial unobserved infections *κ>* 10 are also characteristics of countries will large cumulative confirmed case counts. Conversely, countries that controlled the outbreak and avoided large case numbers during the period of study, such as Australia (AUS), New Zealand (NZL), South Korea (KOR), and Taiwan (TWN) are characterised by either rapid responses, *w_A_ >* 10*^−^*^3^, lower relative initial cases numbers, *κ*, or higher case identification rates *γ*.

Of course, there are exceptions to the trends and changes over time. On 30 March (Fig. 4(A)–(F)), the top ten countries having the largest cumulative case numbers are highlighted in red. In descending order, these are: the United States (USA), Italy (ITA), Spain (ESP), China (CHN), Germany (DEU), France (FRA); Iran (IRN); the United Kingdom (GBR); Switzerland (CHE); and Belgium (BEL). Of these countries, only China was recovering, hence it is highlighted in green. We also highlight South Korea (KOR) as the only other country in recovery during this period, and Taiwan (TWN) as substantial community outbreak was avoided altogether. China and South Korea, countries that recovered rapidly, are characterised by a higher identification rate (China *γ* =0.96; 95% CrI [0.01,0.90]; South Korea *γ* =0.28; 95% CrI [0.17,0.93]) and lower relative initial undocumented cases (China *κ* =0.13; 95% CrI [0.08,35.2]; South Korea *κ* =2.56; 95% CrI [0.50,5.93]) which is indicative of their strict testing, isolation, and tracing regimes [44,45]. Taiwan (TWN), with a high response weight (*w_A_* = 10^−1.9^; 95% CrI [10^−5.9^,10^−1.1^]) responded very rapidly, having established strong public health response mechanisms after the 2003 severe acute respiratory syndrome (SARS) outbreak. This is also reflect in a low level of initial undocumented cases *κ* =6.48 (95% CrI [0.18,21.95]) for Taiwan [46]. This is in stark contrast to Iran (IRN; *κ* = 33.79; 95% CrI [4.7,91.41]) that experience substantial community transmission ahead of the first reported cases. Furthermore, the large response weight for Iran is results in almost no effective reduction in community transmission since *α*_0_ is much larger than *α* (See Supplementary Material), which could represent large gatherings within areas of pilgrimage [15, 47, 48].

By 13 of April, the situation changes as COVID-19 spreads to more countries and response strategies are altered. The top ten countries having the largest cumulative case numbers had changed to be, in descending order: The United States (USA); Spain (ESP); Italy (ITA); France (FRA); Germany (DEU); The United Kingdom (GBR); China (CHN); Iran (IRN); Turkey (TUR); and Belgium (BEL). By this time, Australia (AUS) and New Zealand (NZL) were included in the ranks of countries that were starting to recover. In Fig. 3(D)–(E), there is a substantial decrease in the response weight parameter many of the worst affected countries (USA, *w_A_* = 10^−5.2^ 95% CrI [10^−5.7^,10^−3.2^]; FRA, *w_A_* = 10^−4.7^ 95% CrI [10^−5.4^,10^−1.3^; ESP, *w_A_* = 10^−4.4^ 95% CrI [10^−4.7^,10^−3.9^; DEU, *w_A_* = 10^−4.4^ 95% CrI [10^−4.8^,10^−4.1^; IRN, *w_A_* = 10^−4.4^ 95% CrI [10^−5.2^,10^−1.8^; ITA w*_A_* = 10^−4.2^ 95% CrI [10^−5.8^,10^−1.2^; GBR *w_A_* = 10^−4.0^ 95% CrI [10^−5.1^,10^−1.1^;). This indicates that, in light of data between 31 March–13 April, the community response to the outbreak been even more delayed than earlier data indicated. Conversely, the very high values of *w_A_* for New Zealand (*w_A_* = 10^−2.56^; 95% CrI [10^−5.43^, 10^−1.25^]) demonstrates that a rapid response has been a key factor in keeping cumulative case number low (*C_T_ <* 1, 500). Many countries in the top ten cumulative case numbers still had a larger point estimates for *κ>* 10 (Fig. 4(E),(F)), with the United Kingdom having the largest point estimate of *κ* = 42.73 (95% CrI [0.46,91.52]), which could be the result of early unobserved transmission prior to abandonment of “herd immunity” targets in favour of social distancing, and closing of non-essential business and schools [18,49]. Australia has a very similar characterisation to the United Kingdom in terms of response timing (*w_A_* = 10^−3.36^; 95% CrI [10^−^_3.59,_ 10^−3.22^]) but with much lower *κ* =2.72 (95% CrI [0.24,2.78]) and higher *γ* =0.16 (95% CrI [0.16.0.99]) that likely reflects fact that many of confirmed cases within Australia during this period were imported cases and local community transmission was low [25]. During this time period, small increases in *γ fo*r Germany (*γ* =0.15; 95% CrI [0.01,0.87]), Italy (*γ* =0.10; 95% CrI [0.0,0.92]) and France (*γ* =0.07; 95% CrI [0.01,0.84]), and a large increase in *γ fo*r the United States (*γ* =0.21; 95% CrI [0.0,0.89]); this is possibly a reflection of increased testing capabilities within these countries between 31 March to 13 April [50]. Overall, there is also a decrease in *κ*, indicating that the number of early undocumented infections could be lower than previously thought. Especially for Germany with *κ* =3.27 (95% CrI [0.44,38.63]).

In our final analysis period (Fig. 4(G)–(H)), 9 June, Brazil (BRA), Peru (PER), Russia (RUS) and India (IND) have replaced Belgium (BEL), China (CHN), Iran (IRN) and Turkey (TUR) in top ten countries for cumulative case numbers as the epicentre of the COVID-19 pandemic shifts away from Europe. Once again, these four new countries in the top ten cases list are characterised by low response weights (BRA, *w_A_* = 10^−5.2^ 95% CrI [10^−5.7^,10^−1.1^]; IND, *w_A_* = 10^−4.1^ 95% CrI [10^−5.8^,10^−1.0^]; PER, *w_A_* = 10^−3.8^ 95% CrI [10^−5.6^,10^−1.1^]; RUS, *w_A_* = 10^−4.9^ 95% CrI [10^−5.6^,10^−4.7^]), low identification rates (BRA, *γ* =0.03 95% CrI [0.01,0.90]; IND, *γ* =0.01 95% CrI [0.01,0.88]; PER, *γ* =0.004 95% CrI [0.002,0.83]; RUS, *γ* =0.05 95% CrI [0.02,0.92]) and high relative numbers of initial undocumented infections (BRA, *γ* = 79.34 95% CrI [1.19,80.59]; IND, *γ* = 47.62 95% CrI [0.95,92.97]; PER, *κ* = 15.47 95% CrI [0.80,89.93]; RUS, *κ* = 19.39 95% CrI [0.44,61.41]). At this point in time, many of the mainland European countries were recovering and relaxing restrictions imposed by intervention strategies, consequently the parameters for Germany (DEU), Spain (ESP), France (FRA), and Italy (ITA) have not changed much from the previous analysis other than a further reduction in the estimated relative initial undocumented case numbers for Germany (*κ* =0.11; 95% CrI [0.01,43.80]). While the United Kingdom (GBR) now has higher response weight, this is unfortunately offset by a larger initial relative number of undocumented infections. For the United States (USA), there is a decline in the identification rate (*γ* =0.14 95% CrI [0.01,0.96]).

Overall, our analysis of the first wave identifies features of how communities globally respond to the COVID-19 pandemic without explicit modelling of specific NPIs. Globally, we see that a delay in regulatory response (small *w_A_*) corresponds to countries having high numbers of cumulative case numbers. Fortunately, it seems that many countries had responded rapidly by introducing interventions when the earliest cases where identified. However, many countries also demonstrate large values of *κ*, indicating that delays in early detection have lead to a larger unobserved infectious population. As a result, the active case curve will have taken longer to turn around. In the event of future outbreaks, this downturn could be brought forward by increasing *γ* through increased identification and quarantine/isolation capacity.

### Overall Assessment of the global response to the COVID-19 outbreak

We present an overview of the relationship between model parameter estimates and the magnitude of the COVID-19 outbreak. Here we use the point estimates computed for 121 countries for time period 22 January to 13 April. We use these estimates to evaluate what factors have had the greatest impact on the COVID-19 outbreak evolution across countries. For each country, we manually classify the state of the outbreak for each country on 13 April based on the trend in the daily reported cases and active case numbers.

These stages are: the growth stage–characterised as an increasing trend in daily reported cases numbers; the post-peak stage–characterised by declines in daily case numbers, indicating the curve is flattening; the recovery stage–characterised by declines in active case numbers. Spearman’s rank-order correlation coefficients are computed between each parameters and observed data at *T* = 13 April (e.i., cumulative case numbers, *C_T,_* recoveries, *R*, and deaths, *D_T_*). Figure 5 highlights our results from this analysis. The lower diagonal section of Fig. 5 show the distribution of point estimates and outbreak stage classifications, whereas the upper diagonal show the correlation coefficients.

**Figure 5:**
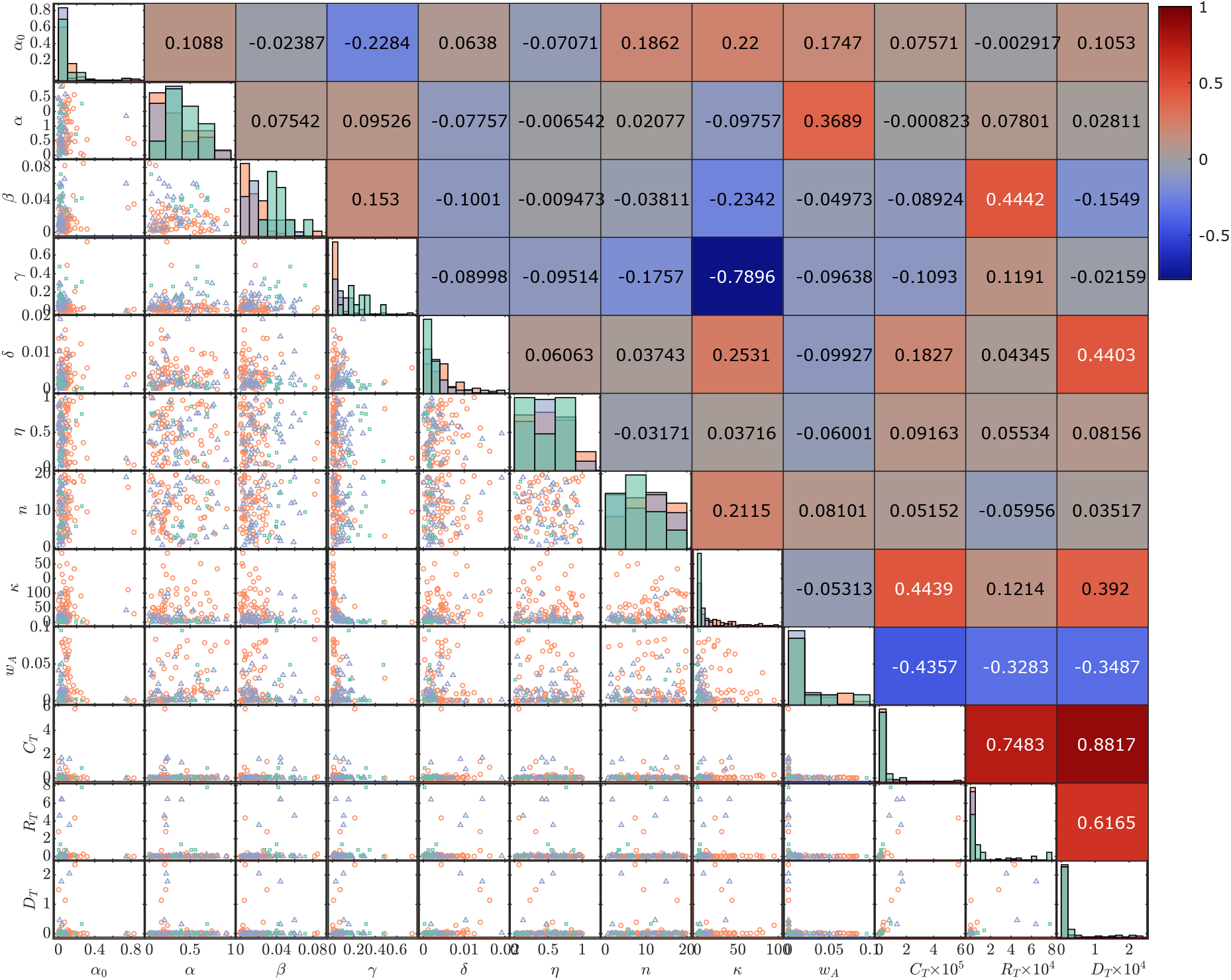
Distributions of model parameter point estimates along with observered cumulative confirmed cases *C_T,_* recoveries *R_T_* and deaths *D_T_* at *T* = 13 April. Pairwise scatter plots on the lower diagonal indicate the stage of the COVID-19 outbreak for that country: growth stage (red circles), post-peak stage (purple triangles), or recovery stage (green squares). Histograms on the diagonal show the distribution of parameters across all countries within each outbreak stage. Spearman correlation coefficients between each point estimate and observed case numbers with the sign and strength of the correlation indicated by the colour-map (positive correlations in red and negative correlations in blue).

The two parameters with the strongest correlation with large case numbers, C*_T,_* are the response weight parameter *w_A_* (Spearman’s *ρ* = −0.4357), and the relative initial undocumented cases numbers, *κ*, (Spearman’s *ρ* =0.4439). These parameters relate to the response behaviour of communities at the early stages of the outbreak. Recall *A_t_* =1*/w_A_* is the critical number of active cases to invoke a response strength of 50%, corresponding to a reduction in transmission rate of 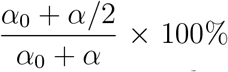. Therefore, a smaller value of *w_A_* indicates a delay in community response, since larger numbers of active cases are required to invoke a response strength of 50%. Similarly, a large values for *κ* are indicative of community transmission occurring ahead of the earliest reported cases. Furthermore, there is a strong negative correlation between the identification rate *γ* and *κ* (Spearman’s *ρ* = −0.7896), indicating that countries with strong testing and contact tracing regimes were able to minimise the amount of undocumented community transmission at early time. The identification rate also has low negative correlation (Spearman’s *ρ* = −0.2284) with the residual transmission rate, *α*_0_, meaning that countries with stronger testing regimes also improved maximum efficacy of other interventions. Interestingly, the response slope parameter *n* had only weak correlations with any other parameters. This parameter relates to the rate of change in community behaviour was before and after the critical *A_t_* =1*/w_A_* point (See Fig. 2 in Methods). Note the weak positive correlation between *κ* and *n*, meaning that countries that adopted a more gradual introduction of interventions (lower *n*) also had less community transmission occurring. While the three outbreak classification stages are not well separated in all parameters, there are a few trends to note. Countries that are in the recovery stage tend to have lower residual transmission, *α*_0_, larger regulated transmission, *α*, larger case recovery and identification rates, *β* and *γ*, lower death rates *δ*, and lower relative undocumented initial infections *κ*. For countries still in the growth stage of the outbreak, the converse is true. Unsurprisingly, the post-peak stage (i.e., declining daily cases, without decline in active cases) have, on average, parameter values that sit between the recovery and growth stages. It is important to note some countries that experienced large numbers of cases in this analysis period are also in the post-peak or recovery stage, whereas others in the growth stage had only small numbers of cases at this time. In some case, such as countries of South America, this analysis could have helped highlight the importance of interventions at early stages of the pandemic.

### Avoiding second waves

As societies begin to return to normal, reliably estimating the number of undocumented infections, including the extent of asymptomatic by infectious case, is crucial to avoid flare-ups that potentially can lead to second waves of COVID-19 spread [51]. Recent evidence suggests asymptomatic individuals having a substantial role in the spread of COVID-19 [52]. Here, we demonstrate that quantification of uncertainty in the unobserved infectious population is crucial for planning the timing of easing of restrictions.

Due to the form of our response function in our model, we can model both how communities introduced and subsequently relax interventions as active case numbers reduce. That is, as *A_t_* declines then *f*(·) increases to simulate increase mixing of the population. The posterior predictive distribution can start to demonstrate oscillatory behaviours, the magnitude of which depends on the evolution of the undocumented infectious population *I_t_*. For example, Fig. 6 demonstrates this behaviour by extending simulations up to the 24 June. In both these examples, uncertainty in the daily case numbers, driven by parameter uncertainty and undocumented infectious population uncertainty, pre-empts the small flare-ups in numbers. To obtain this behaviour, all possible evolutions of I*_t_* that are consistent with the observed daily cases must be taken into account.

**Figure 6:**
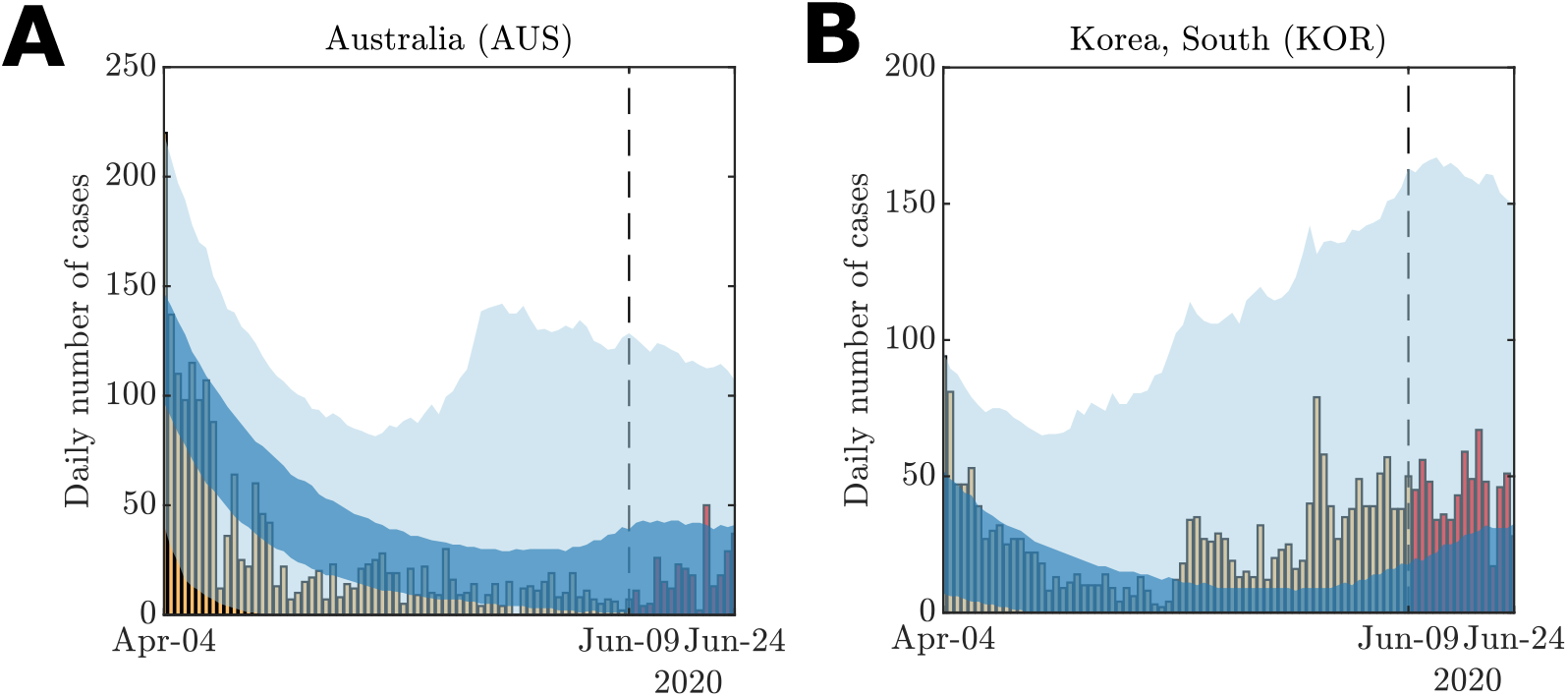
Example of small secondary oscillations in model behaviour using the model fit against daily case data (yellow bars) for (A) Australia and (B) South Korea up to 9 June (dashed line). The posterior predictive simulations are continues up to 24 June to demonstrate the uncertainty in potential case increases after relaxation of restrictions. Actual daily case numbers for the period 10–24 June (red bars) also demonstrate increases within the credible intervals (dark blue 50% CrI; light blue 95% CrI).

The key message from this analysis is to highlight the importance of conservative uncertainties in the future evolution of the pandemic. It is encouraging to see predictions that indicate potential future declines in active cases counts. However, uncertainties in the number of undocumented cases and model parameters indicate caution in predicting the timing of consistent declines in active case numbers. Therefore, it is essential that communities remain vigilant in fast-evolving situations such as this.

## Discussion

We have applied a novel stochastic epidemiological model to characterise the response to the first wave of the COVID-19 pandemic. We find that the worst affected countries (in terms of confirmed case numbers), are characterised by a delayed response (small *w_A_*), allowing case numbers to rise before interventions became effective. However, increased testing and isolation protocols (large *γ*) have demonstrably reduced the longer term impact, as demonstrated by China and South Korea. Many countries seem to be learning from these collective experiences, with more rapid responses (large *w_A_*). Unfortunately, we also identify that the number of undocumented cases likely substantially exceeded the confirmed cases for many countries (large *κ*). It is important to emphasise that we do not make any specific guidelines for particular countries in improving specific non-pharmaceutical intervention (NPI) or testing strategies. However, in light of our analysis, we advise that intervention mechanisms be mobilised rapidly without waiting for large numbers of cases to be confirmed. This has been a key characteristic of countries that have successfully managed the initial outbreak, such as Taiwan and New Zealand.

The data on reported daily cases from the Johns Hopkins University coronavirus resource center have some limitations [12]. Firstly, when aggregated at a country level, these data do not take into account high levels of spatial heterogeneity. To account for potential bias, future work could consider a sensitivity analysis on the level of individual cities or provinces where available. The delay between onset dates and reported dates for new case also potentially introduces bias and data spikes (as noted in Methods and Results). Lastly, the Johns Hopkins data does is not curated to distinguish between cases acquired through local transmission as opposed to imported cases. Further modelling extensions that account for details captured in alternative data sources, such as the European Center for Disease Control and Prevention (ECDC) [13], should also be considered.

Our model, like any model, has fundamental assumptions that are necessarily introduced. We treat each country as a single well-mixed population. While our approach does include important features such as undocumented infections and a variable transmission rate, more advanced analysis could be performed by considering disease spread through a network [17, 53–55] of well-mixed populations, such as provinces, states or cities. This would assist is capturing social factors that could also influence COVID-19 transmission, such as spatial variation in population density and large population movement such as those that occur during times of festival [14, 47]. We also treat each country as a closed system, whereas realistic sources and sinks through inclusion of an international travel network could enable us to track the impact of decisions of one country on connected countries.

Other details of the model could also be extended. We treat active confirmed cases as non-infectious due to quarantine and isolation. In reality, active confirmed cases can still spread the virus to medical staff. We also apply the reasonable approximation that there is no re-detection or re-infection, however, new evidence is questioning the validity of this assumption [49, 52, 56]. It is also possible that seasonal effects related to climate could also cause transmission rates to change, although the evidence suggests that this is not a substantial effect in the pandemic stage [57]. A finer granularity of classes of susceptible individuals (i.e., at risk), incubation periods, severity of symptoms, and climate effects would also enable more detailed analysis for an individual country [25,28]. However, our reduced set of classes and interactions represents a trade-off between realism and broad applicability to worldwide data. A Bayesian hierarchical modelling approach could also be applied to better capture heterogeneity across countries.

Our model framework is flexible through the inclusion of a response function (See Methods) in the virus transmission mechanism and may be extended to other scenarios. In this manuscript, we have only considered the case of a response dependent on the number of active confirmed cases, leaving a sensitivity analysis for the more general from for the Supplementary Material. However, this response function could be further extended to include economical factors or be modified to be a function of state and time. This would enable a wide range of behaviours to be explored, such as specific timings of enforced NPIs, and subsequent lifting of restrictions when active case go below a threshold.

## Conclusions

Our work confirms that a multi-pronged approach to combat COVID-19 is essential. Firstly, early introduction of testing and effective contact tracing protocols and quarantine effectively reduces the uncertainty in the unobserved infected population (i.e., low *κ an*d high *γ*). Intervention strategies are also essential and are most effective when introduced early (high w*_A_*). These results demonstrate the utility of modelling combined with high quality, immediately available data for providing insight into the eary stages of the pandemic.

It is hoped that our work might be used to inform future responses to outbreaks of COVID-19 or other pandemics. The message is clear: to avoid second waves we must not be complacent in response to an outbreak as the earliest confirmed cases arise. We also highlight the importance of wider testing to effectively reduce uncertainty in predictions of case numbers, recoveries and deaths.

### Ethics approval and consent to participate

Not applicable.

### Consent to publish

Not applicable.

### Availability of data and materials

Data and analysis code used in this study are available on GitHub https://github.com/davidwarne/covid19-auto-reg-model. Data may also be acquired from the Johns Hopkins University coronavirus data repository https://github.com/CSSEGISandData/COVID-19.

### Funding

AE is supported by a SNF grant (105218 163196). CD is supported by an ARC Discovery Project (DP200102101). KM is supported by an ARC Laureate grant (LF150100150). This project has been supported by the Swiss Data Science Center (SDSC, Project BISTOM C17-12).

### Author’s contributions

AM, KM and DJW designed the research. AM, KM, CD and DJW provided analytical tools. AE, CD, and DJW contributed computational tools. AE and DJW processed data. AE and DJW performed simulations and inference. DJW performed the analysis. AM, KM, WH, a DJW interpreted the analysis results. AM, KM, WH, CD, AE, and DJW wrote the paper.

## Data Availability

Data is all available in the public domain.

https://github.com/CSSEGISandData/COVID-19

## Acknowledgements

DJW, KM and CD are members of the Australian Research Council (ARC) Centre of Excellence for Mathematical and Statistical Frontiers (ACEMS). Computational resources were provided by the eResearch Office, Queensland University of Technology.

